# Identifying variation in GP referral rates: an observational study of outpatient headache referrals

**DOI:** 10.1101/2022.02.07.22270572

**Authors:** Fran Biggin, Quinta Davies, Timothy Howcroft, Hedley Emsley, Jo Knight

## Abstract

**Objective:** To identify GP surgeries with unexpected rates of referral to specialist services, using headache referrals to outpatient neurology as an example. Identifying surgeries with unexpectedly high or low referral rates allows for further investigation and potential support to be targeted where it is most likely to be effective.

**Methods:** This is a retrospective observational study using routinely collected and open-source data. Data was collected from a single consultant outpatient neurology clinic and 202 GP surgeries across seven CCGs in the Northwest of England. The number of headache referrals from each GP surgery during a study period of 3 ¼ years was used as the primary outcome in a poisson model. The standardised residuals from this model were then used to identify GP surgeries that were likely to have referred unexpected patient numbers for headaches to an outpatient neurology clinic during the study period.

**Results:** We identified four GP surgeries with unexpected numbers of referrals. This model also showed that there were two main predictors of headache referral, namely other neurology referrals and the distance of the GP surgery from the outpatient clinic.

**Conclusion:** GP surgeries with unexpected numbers of referrals to specialist services were identified using a flexible methodology. This methodology was demonstrated using headache referrals but could be adapted to any type of referral or geographical area.

## Introduction

### Referral

General Practitioners (GPs) provide a number of key services, including referral to specialist treatment when needed. Referrals can be made for a number of reasons including for investigation, diagnosis, management or reassurance [1]. In the UK referral rates from GPs vary for a number of complex reasons such as resource availability, population health needs, patient pressure, and lack of consensus on which conditions benefit most from specialist input [1, 2].

Variability in referral rates from GPs to specialist services is a complex issue with interacting social, geographic, and demographic influences. Understanding variability in referral rates has been an area of interest in health research for many years. In 1989 Coulter et al [3] found that there were many reasons given for GP referral, including to establish diagnosis, for a test or investigation, for treatment, for advice on management, and to reassure both the GP and/or the patient. These differing reasons for referral can contribute to the observed variations in referral rates [4]. Other research has shown that individual GP characteristics such as the ability to tolerate risk also affects rates of referral [5, 6]. A literature review conducted in 2000 identified 91 relevant papers and from these summarised the amount of variation found in referral rates and identified reasons for this variation [7]. It found that most variation in referral rates was unexplained, with patient and GP characteristics only accounting for half of all observed variation.

A number of different research approaches have been made to investigate referral rates, two recent studies have used poisson regression to investigate referrals from primary care to specialist services. Jessen et al [8] used a poisson model to investigate the relationship between GP cancer suspicion and referral rates to standardised cancer referral pathways. They found that referral rates varied by cancer type and whether a GP had an initial suspicion of cancer. Kaur et al [9] used poisson regression to investigate referrals for physical therapy for osteoarthritis during the COVID-19 pandemic.

Over the years other work has focused on investigating interventions that may reduce unnecessary variations in referral and has come to contradictory conclusions. Fertig et al [10] concluded that ‘inappropriate referrals’ were not the cause of variation, and that guidelines may therefore not reduce referral numbers. However, there have been further studies since, including a systematic review by Akbari et al [11] who found effective interventions included targeted dissemination of guidelines and involvement of consultants in educational activities for GPs.

Recently the National Health Service (NHS) has tried to identify potential areas for improvement, both in terms of patient outcomes and reducing costs. Two initiatives include RightCare and Getting It Right First Time (GIRFT). GIRFT aims to ‘improve medical care within the NHS by reducing unwarranted variations’ [12]. The GIRFT Neurology project divides England into ‘neuroscience regions’ for analysis and examines visits to NHS Trusts to ‘deep dive’ into local issues [13, 14]. The NHS RightCare initiative seeks to help CCGs ‘Diagnose the issues and identify the opportunities with data, evidence and intelligence; develop solutions, guidance and innovation; and deliver improvements for patients, populations and systems’ [15]. However, the RightCare methodology has been criticised, including the manner in which similar CCGs are identified, and the way in which CCGs are compared, resulting in overestimation of differences [16]. The methodology we develop in this paper to identify GP surgeries of interest is not intended as a replacement for either RightCare or GIRFT methodology, but offers an alternative approach.

### Headache Referral

In this study we focus on the specific issue of referrals from primary care to neurology outpatient care for headaches (including migraine).

Headache is a common and disabling condition, with migraine representing the second largest contribution to global disability of all neurological conditions [17]. Headache, including migraine, accounts for a large proportion of consultant neurologist appointments in the UK [18–22], and is a common presenting complaint at GP surgeries. GPs refer between 2 to 3% of the headache patients they see in primary care [23], and report experiencing pressure from patients to refer to specialist care [24], despite evidence that headache conditions are often best managed at primary care level [25]. This puts pressure on both GP and outpatient neurology services [26].

GPs are under pressure to provide quality referrals to specialist care, including neurological outpatient care. Brilla et al (2008) found that interventions made at the neurology service level for reducing ‘inappropriate referrals’, such as email triage, are ineffective [27]. This study concluded instead that interventions should be made at the point of referral by enhancing guidance for referral decisions for GPs. Davies et al also emphasised the benefit of interventions at primary care level, recommending improved education for GPs to help reduce the burden of headache [28]. Most recently Huang et al found that an online headache referral guideline for GPs was successful in reducing the number of referrals to neurology services [29].

If interventions such as structured guidelines and education are best applied at the point of referral, then identifying where these interventions may be most effective would be of interest. In addition, identifying GP surgeries where referral rates are reduced may offer further insights into the spectrum of variation in referral patterns, as under-referral can also potentially signal the need for intervention to improve care.

### Specific Objectives

In this study we identify GP surgeries with unexpected rates of referral. Although the specific case study used is headache referrals, this study aims to provide a methodology which is flexible and can be applied to any type of referral both within and outside of the neurology specialty.

## Methods

### Study Design

We used routinely collected data from outpatient appointments alongside open access data in a retrospective observational study. We recorded the number of patients referred for headache to, and offered an appointment in, a single consultant-delivered neurology clinic over a period of three years and four months (18^th^ September 2015 to 9^th^ January 2019). We had access to indentifiable information during data collection, but data were anonymised before analysis.

The study received relevant approvals, including NHS Research Ethics (Ref: 19/NW/0178) and Confidentiality Advisory Group (Ref: 19/CAG/0056), as well as Health Research Authority (HRA) on 30 May 2019 (Ref: 255676). The study was also approved by the Lancaster University Faculty of Health and Medicine Research Ethics Committee on 17 June 2019 (Ref: FHMREC18092).

### Data Sources

Data regarding the number of referrals from GPs within the catchment CCGs were taken from neurology outpatient clinic records at the Royal Preston Hospital (RPH), which is part of the Lancashire Teaching Hospitals NHS Foundation Trust (LTHTR). As the clinic is dedicated to adult care no paediatric referrals were included. The data covers all GPs within 7 Clinical Commissioning Groups (CCGs): Greater Preston; Chorley and South Ribble; East Lancashire; Fylde and Wyre; Blackpool; Blackburn with Darwen; and Lancashire North. A small number of referrals for headache arose outside this catchment area, but their small number made them unsuitable for inclusion in the analysis (6 from Cumbria CCG, and 1 each from West Lancashire and Wigan Borough CCGs).

Data regarding GP surgery characteristics was downloaded from NHS Digital open access repositories https://digital.nhs.uk/services/organisation-data-service/data-downloads/gp-and-gp-practice-related-data This study did not use the latest data available as, although the data is updated regularly, we felt it more appropriate to use data from the start of our study period (October 2015). This allowed us to capture information for GP surgeries that have subsequently been closed or amalgamated with other locations.

### Variables

#### GP surgery variables

The outcome of interest was the number of headache referrals from each GP surgery during the study period. Explanatory variables were chosen for both their relevance and their availability. These variables included GP surgery list size (adults over 14 years of age); proportion of males; mean age; number of other neurology referrals made; distance of the surgery from the clinic at Royal Preston Hospital (RPH); weighted Index of Multiple Deprivation (IMD); and the standard deviation of the weighted IMD. Previous studies have found that socioeconomic deprivation, young age, and female gender appear to be associated with greater headache burden and the likelihood of referral [1, 30, 31], hence the inclusion of IMD, age, and gender in our analysis.

#### Weighted IMD calculation

The Index of Multiple Deprivation (IMD) is often used as an indicator of the deprivation of an area. It is collected at the census unit of the Lower Super Output Area (LSOA). As GP catchments overlap fragments of many LSOAs, to be able to explore relative deprivation levels of GP surgeries we calculated a weighted score for each surgery. We followed the methodology devised by Zheng et al. [32]. We combined data on the number of patients from each LSOA on each GP’s list (available online from NHS Digital https://digital.nhs.uk/data-and-information/publications/statistical/patients-registered-at-a-gp-practice/october-2015) with IMD data available from the gov.uk website (https://www.gov.uk/government/statistics/english-indices-of-deprivation-2015), creating a weighted index for each GP surgery. We also created a variable for the standard deviation of those weighted indices as a measure of the variability of the IMDs contributing to each GP surgery.

### Statistical Methods

#### Data Preparation

The data were analysed using R Studio version 1.2.5019. The separate datasets were joined using GP surgery codes. After joining, we calculated weighted IMD, standard deviation of the weighted IMD, and the straight-line distance of each surgery from the clinic at RPH using the Ordnance Survey coordinate system.

#### Modelling

First we identified which variables drive variation in referral numbers and then calculated expected referral rates from the GP surgeries using a poisson log-linear model with an offset for GP surgery list size. We included the CCGs as a factor with Greater Preston CCG as the comparator. We included list size as an offset in order to include both list size and ‘other neurology referrals’ as potential factors influencing referral. We chose a poisson model as it provided the best fit when compared with zero-inflated poisson, negative binomial, and zero inflated negative binomial. Models were compared using the Akaike Information Criterion (AIC).

Our model included data from GPs in the 7 CCGs which make up the catchment area for the clinic. Towards the outer edges of this area it is likely that some patients are referred elsewhere for neurology outpatient care. GPs and patients have an element of choice of outpatient clinic to which to refer/visit, and one of the factors influencing this choice is likely to be distance from the clinic. In general, the farther the GP surgery is from the clinic at RPH, the more likely a patient is to choose an appointment elsewhere. We account for this by including both distance from the clinic at RPH and the number of other types of neurology referral made by each GP surgery.

#### Examining the differences

In order to identify surgeries which refer fewer or more patients that expected, we extracted predicted values from the model described above and compared them to the actual values observed during the study period. This gave us the differences between observed and expected referrals which can be visualised to determine if surgeries are referring as expected. We also examined potential spatial autocorrelation between the differences using Moran’s I. This index is similar in concept to a correlation coefficient and gives a value between −1 and 1. However, −1 indicates perfect clustering of dissimilar values, and 1 perfect clustering of similar values. A Moran’s I of 0 indicates perfect randomness. Investigating spatial autocorrelation allows us to determine if GP surgeries which are located close together are more likely to have similar differences between observed and expected number of referrals than those further apart, and thus if there are any potential factors related to location which affect referrals.

## Results

We analysed data from 202 GP surgeries across 7 CCGs (see Fig 1), which provided 388 headache referrals in total over the period of the study. Over the same period these surgeries referred 1371 patients for other suspected neurological disorders, thus headache accounts for 19% of all neurology referrals over this study period. The largest number of surgeries is in East Lancashire CCG, but the largest number of both headache and other neurology referrals was from the Greater Preston CCG. This may stem from the fact that GPs can refer patients to other neurology clinics if the patient prefers, and the farther away a GP surgery is from the outpatient clinic, the more likely they are to refer elsewhere. The characteristics of the GP surgeries in each CCG can be found in Table 1.

**Fig 1.**
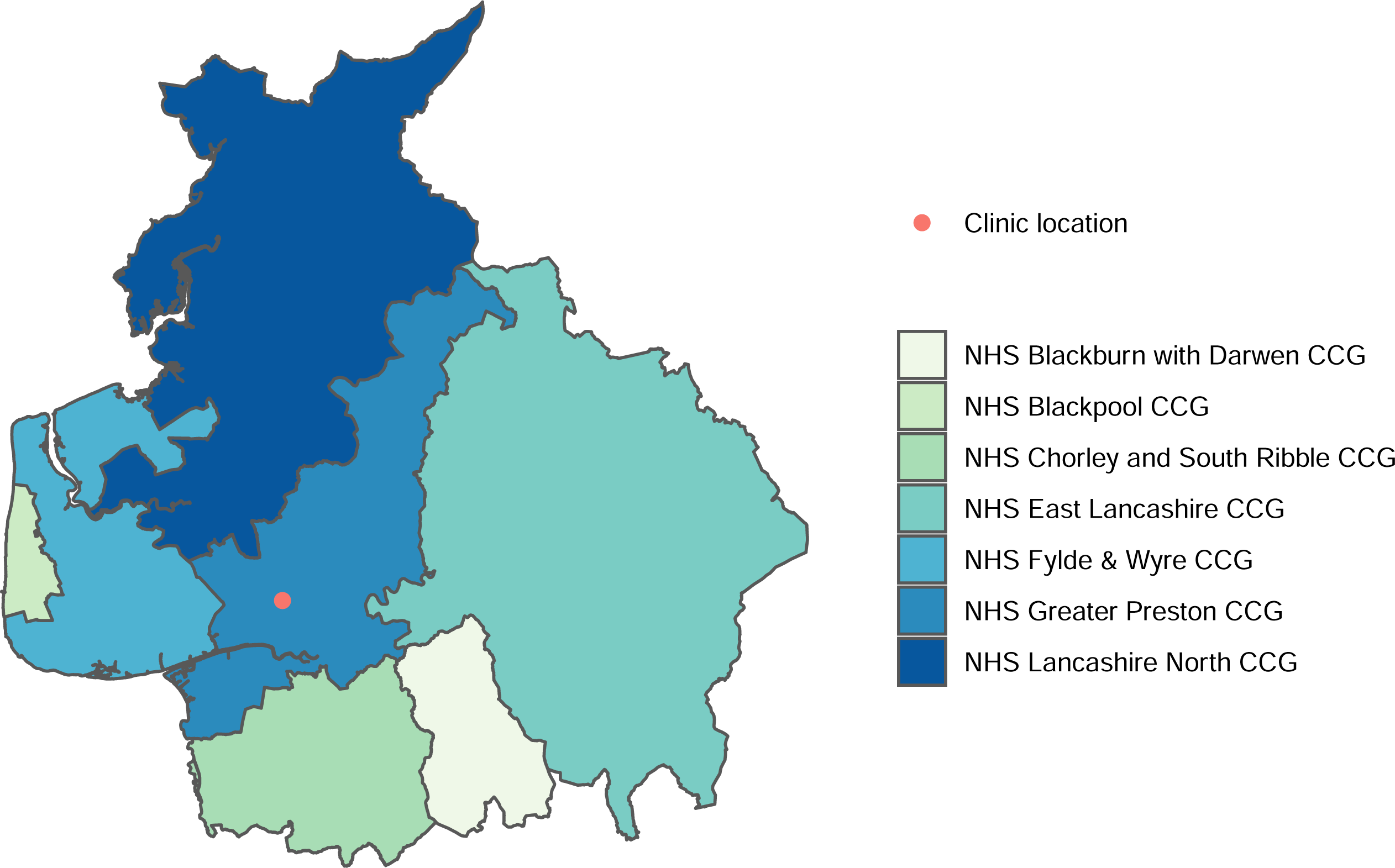
A map of the contiguous CCGs included in the study.

**Table 1.** Study Characteristics.

| Clinical Commissioning Group | Number of Surgeries | Number of Headache Referrals | Number of other neurology referrals | GP Surgery Characteristics |  |  |  |  |
| --- | --- | --- | --- | --- | --- | --- | --- | --- |
|  |  |  |  | Mean Age of List (range)* | Mean List Size (range)* | Mean Weighted IMD (range)* | Mean Distance from RPH in km (range)* | Mean Percentage of Males (range)* |
| Blackburn with Darwen | 27 | 26 | 110 | 43 (37-47) | 5020 (1303-13273) | 3.4 (1.5-5.9) | 16 (14-19) | 51 (47-55) |
| Blackpool | 22 | 47 | 126 | 46 (39-52) | 6577 (1939-12525) | 2.8 (1.4-4.6) | 22 (21-24) | 51 (48-54) |
| Chorley and South Ribble | 31 | 61 | 237 | 46 (38-51) | 4695 (1190-13651) | 6.4 (3.7-9.1) | 13 (6-21) | 50 (47-54) |
| East Lancashire | 57 | 53 | 206 | 45 (36-50) | 5267 (894-15839) | 3.8 (1.4-8.5) | 28 (20-40) | 51 (47-55) |
| Fylde and Wyre | 21 | 65 | 123 | 49 (45-53) | 6442 (1507-10167) | 5.8 (2.7-7.5) | 20 (11-25) | 49 (47-53) |
| Greater Preston | 32 | 111 | 503 | 44 (35-49) | 5214 (1427-14408) | 4.5 (1.6-9.1) | 4 (0.5-9) | 52 (48-60) |
| Lancashire North | 12 | 25 | 66 | 45 (31-50) | 10768 (5824-26512) | 5.5 (3.4-7.8) | 28 (13-38) | 50 (47-52) |
| Overall | 202 | 388 | 1371 | 45 (31-53) | 5729 (894-26512) | 4.4 (1.4-9.1) | 19 (0.5-40) | 51 (47-60) |

\* Indicates the range of values from the individual GP Surgeries within the CCGs

GP surgery size varies greatly across the catchment area, from the smallest surgery of 894 patients in East Lancashire CCG to the largest surgery with 26,512 patients in Lancashire North. The distance of the surgeries from the clinic at RPH varies from 550m to 39.5km, with an average distance of 18.9km. The calculated weighted IMD for each surgery varies greatly from a low of 1.4 in Blackpool and East Lancashire CCGs to 9.1 in Greater Preston and Chorley and South Ribble CCGs, indicating marked diversity in the socioeconomic characteristics across the CCGs.

Within the CCGs GP surgeries referred differing numbers of patients for both headaches and other types of neurological conditions. Fig 2 shows the relationship between the number of headache patients a GP surgery referred and the number of other neurology referrals. This relationship is shown separately for each CCG and demonstrates that there is a consistently positive relationship between headache and other neurology referrals, although this relationship appears to differ between CCGs.

**Fig 2.**
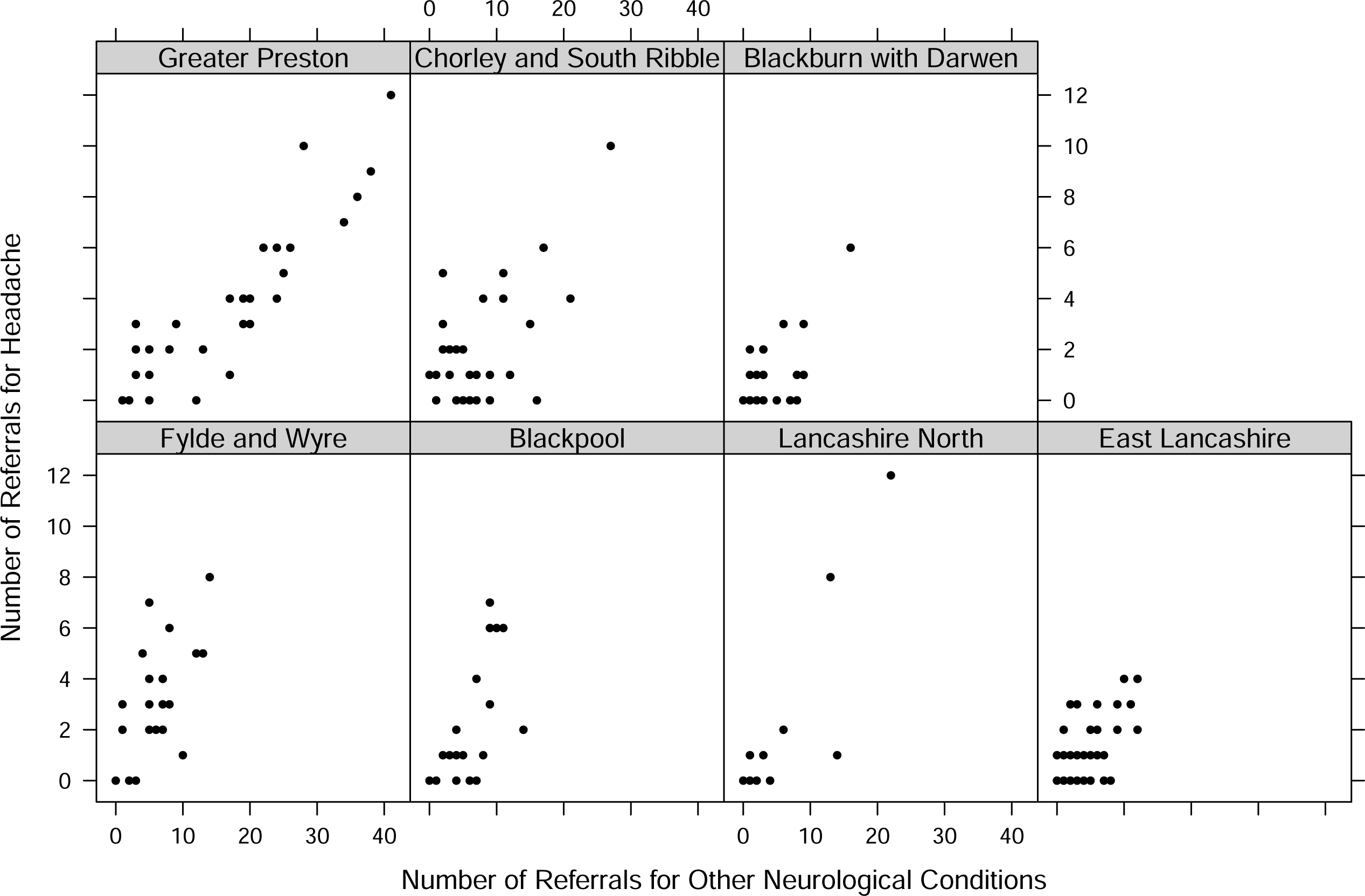
Correlation of the number of headache referrals and other neurology referrals from all GP surgeries split by CCG.

### Model Results

Results from the model can be seen in Table 2. We see variation in referral is influenced by the number of other neurology referrals, and by the distance of a GP surgery from the clinic. These results suggest that the further a surgery is from the clinic the fewer headache referrals are made.

**Table 2.** Results from the poisson model.

|  | Estimate | Std Error | IRR* (95%CI) | P value |
| --- | --- | --- | --- | --- |
| Intercept | -8.06 | 2.72 |  | 0.003 |
| Other Neurology Referrals | 0.04 | 0.008 | 1.04 (1.02-1.06) | < 0.001 |
| Mean Age | 0.03 | 0.02 | 1.03 (0.99-1.06) | 0.21 |
| Weighted IMD | -0.03 | 0.04 | 0.96 (0.88-1.05) | 0.46 |
| SD of weighted IMD | -0.25 | 0.12 | 0.78 (0.61-0.98) | 0.03 |
| Distance from clinic | -4.2x10<sup>-5</sup> | 1.3x10<sup>-5</sup> | 0.99 (0.99-0.99) | < 0.001 |
| Proportion male | -0.78 | 4.3 | 0.46 (7.2x10<sup>-5</sup>-213) | 0.85 |
| Blackburn with Darwen | 0.04 | 0.29 | 1.05 (0.59-1.82) | 0.87 |
| Blackpool CCG | 0.46 | 0.32 | 1.59 (0.85-2.95) | 0.14 |
| Chorley and South Ribble CCG | 0.37 | 0.21 | 1.45 (0.95-2.18) | 0.08 |
| East Lancashire CCG | 0.37 | 0.34 | 1.45 (0.74-2.82) | 0.27 |
| Fylde and Wyre CCG | 0.81 | 0.27 | 2.25 (1.30-3.89) | 0.004 |
| Lancashire North CCG | 0.27 | 0.34 | 1.31 (0.65-2.55) | 0.43 |

\*IRR: Incident Rate Ratio. As the list size of each practice was included as an offset in the model, the output of the model is a rate which depends on the list size of the GP.

### Examining the differences

The standardised difference between the number of expected and observed referrals for each surgery within the seven CCGs is shown graphically in Fig 4. This figure shows a boxplot of the distribution of the overall differences, and a dot for each GP surgery colour coded by CCG. Values below −3 or above +3 can be considered statistically significant.

There were no surgeries which referred fewer headache patients than expected, and 4 surgeries that referred statistically significantly more than expected, given the variables that were accounted for in the model. A plot comparing the raw (non-standardised) values of observed and predicted referrals can be found in the supporting information Fig S1.

Testing for spatial correlation in the differences between expected and observed referrals using Moran’s I, we found there to be no spatial autocorrelation for the GPs across the 7 included CCGs (Moran’s I = −0.013, p = 0.605).

## Discussion

### Principal findings

This study shows that it is possible to identify GP surgeries which refer unexpected numbers of patients to an outpatient clinic. This is achieved by identifying a set of explanatory variables to be included in a poisson model, the results of which are then used to give predicted values for comparison against observed referral numbers. Once surgeries with unusual numbers of referrals are identified, further investigation can then be carried out to understand the circumstances leading to the unexpected referral numbers. This would allow support to be targeted to the places that need it, and lessons to be learnt, which could be shared across the CCGs.

### Strengths and limitations

The basic methodology outlined in this paper could be modified and extended to other specialties. Although other specialties have their own drivers for referral patterns, they could be examined using the same methodology by adjusting the explanatory variables included in the initial model. The methodology could also be extended to cover larger geographical areas.

As with all studies which include statistical modelling, if an informative variable has been excluded, either through unavailability of data or through not understanding the drivers of referral, then the results of the second stage of the process - identification of the unexpected referral rates – would be less accurate. Researchers need to understand the drivers behind the type of referral under investigation, and to be able to access valid data on which to build models. If understanding is limited, or if data is unavailable (or inconsistent) any assumptions drawn from modelling will be flawed.

The way that data is collected from GP surgeries can change over time. Surgeries can be closed, and new surgeries can be created from both amalgamation and splitting of previous surgeries. In this study we extracted data from the NHS Digital open access repositories for the dates at the start of the study as this allowed us to capture information for GP surgeries that have subsequently been closed or amalgamated. In addition, some GP surgeries are small single locations, whereas others comprise of a large hub surgery and several smaller afflilated branch surgeries. In this study we did not split branch surgeries from their parent location, as branches which come under a single surgery grouping are likely to have much in common. For example, they are likely to share the same educational training and use the same referral guidelines.

This study has a relatively small sample size, in particular there are few surgeries which refer large numbers of headache patients. Therefore it should be replicated with a larger dataset, in order to corroborate the results seen in this study. Expanding the study to a larger geographical area, including other clinics, or including a longer study period would also help to alleviate the limitation of a small dataset, as well as expanding the generalisability to other areas. However, this expansion would be reliant on the availability of coded outpatient neurology diagnoses.

The methodology used in this study can be adapted to any type of referral, geographical location, and timescale by adjusting the explanatory variables used in the initial model. Although the results of the model used as a case study in this paper are not generalisable to other geographical locations or timescales, the methodology is generalisable. It would be possible to expand this analysis of headache referrals to a national level, but this would rely on the availability of consistent coding of neurology outpatient appointments.

### Relation to previous studies

Previous studies have used statistical modelling to investigate the impact of different variables on referral rates from GPs to specialist services [8, 9]. We have based the first stage of our study on this modelling process, and then extended the analysis to include the identification of GP surgeries which are referring unexpected numbers of patients for headache.

The NHS RightCare methodology identifies areas of opportunity for improvement for CCGs, and although our study does not seek to replicate or replace the RightCare methodology, the intention behind it is similar – to allow CCGs to identify GP surgeries where interventions may be of use. However our methodology differs in a number of ways from RightCare which allows it to avoid the difficulties highlighted by Dropkin [16]. RightCare compares CCGs across large dislocated geographical distances, whereas we limit the study to a single contiguous geographical area, meaning that the CCGs are more likely to be similar in unmeasured ways. In addition the RightCare methodology has a fixed set of demographic variables against which the CCGs are measured regardless of the outcome of interest, whereas we recommend that the explanatory variables used in the initial model are changed depending upon the outcome under investigation. It cannot be assumed that the explanatory variables used for headache referral would be relevant for orthopaedic referral, for example.

### Meaning of the study

In a previous study we found that the majority of patients with headache who were referred to a neurology outpatient clinic had only one appointment [21]. Many of these patients were discharged after only one appointment without investigation. Whilst a single consultation with a neurologist can make an important contribution to the patient’s management, much of the advice given, particularly in relation to lifestyle factors and avoidance of medication overuse headache, could be delivered in primary care [25]. Demand is rising and capacity, including in general practice, is limited. Identifying which surgeries could potentially support patients through alternative routes to treatment would be both more convenient for the patient and free up resources for other patients needing to access care.

This study provides an indication of GP surgeries from which there may be unexpected numbers of referrals, but it does not explain why those unexpected referrals may have occurred. However, identifying variation is the first step towards understanding it, and this methodology could be used by CCGs or outpatient clinics to understand where their patients are coming from, and to plan further targeted investigations.

### Unanswered questions and future research

More research needs to be done to validate this methodology with a larger dataset, and to extend it into other areas of referral. It could also be extended and refined to apply to larger geographies, or to other outpatient specialties. Expanding this research to larger geographical areas would require consistent coding of diagnoses resulting from outpatient neurology appointments, which is unfortunately not yet available. Further research is also needed to confirm the utility of conducting these types of analysis, in particular whether identifying unexpected referral rates leads to implementation of policies that improve patient care.

It would also be of interest to analyse what happens to patients following referral in order to determine if the referral was ‘appropriate’ or useful to the patient. This would necessitate collecting qualitative data on the patient experience and would provide a more holistic view of referrals. Another area that we were unable to explore in this paper is any alternative treatment that a patient may seek if denied the opportunity for a referral to specialist consultant care. Future research could address this by examining whether patients who attend the GP with symptoms of headache and are not refered to specialist care are more likely to attend more appointments with the GP, or seek treatment elsewhere such as at the emergency department.

## Conclusion

Identifying GP surgeries with unexpected numbers of referrals is a useful first step towards understanding the larger issue of variability in referral rates. Once identified, those GP surgeries with unexpected numbers of referral can be investigated further to help understand why their referral rates differ from those expected, and if necessary, interventions can be targeted to where they are most needed. Using GLMs is an efficient way of including explanatory variables that are relevant to the type of referrals under investigation and variables can be changed to directly relate to any type of referral requiring investigation. This ensures that the methodology presented here is flexible enough to be applied to different types of referral or geographical area.

**Fig 3.**
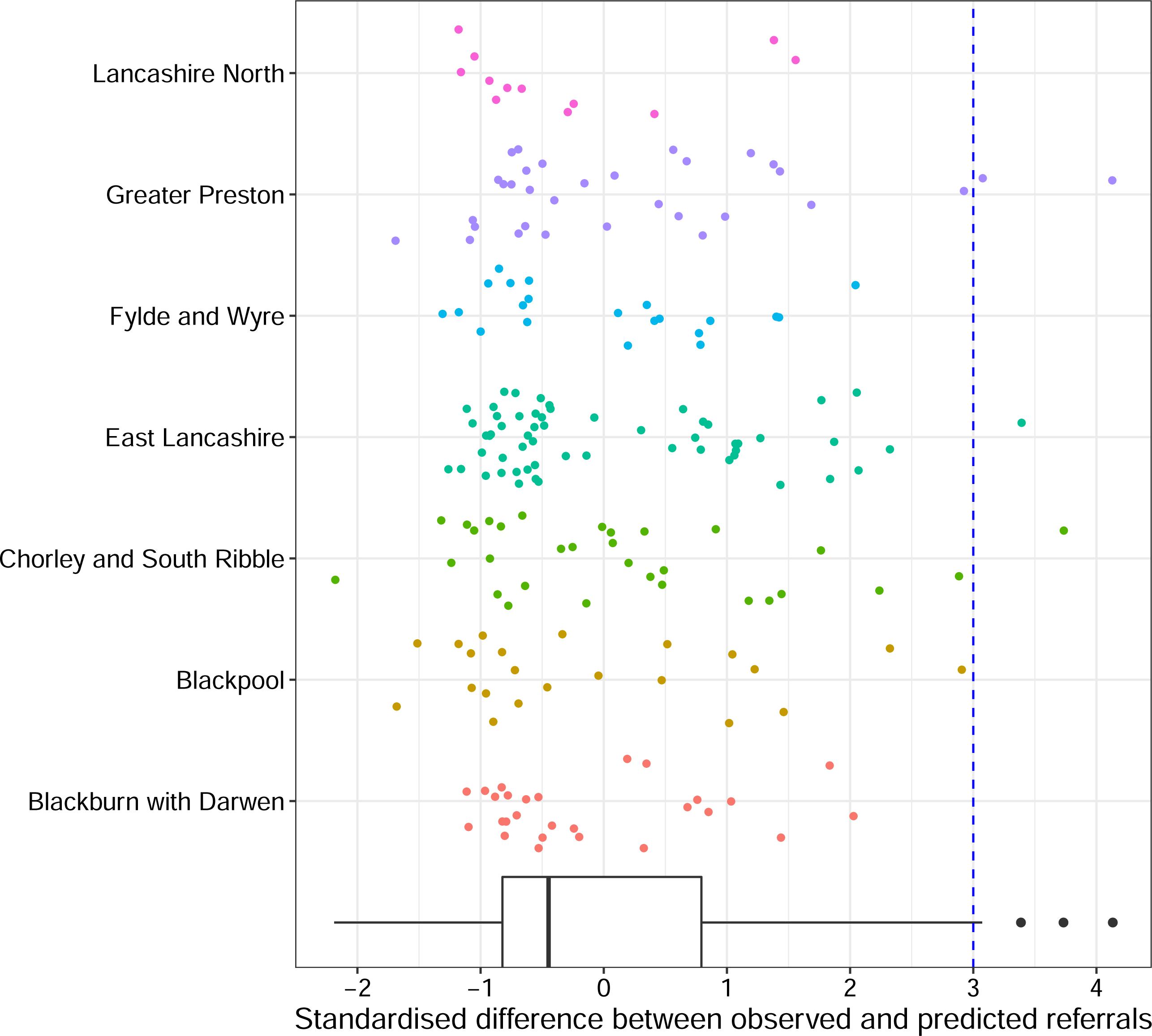
Boxplot and point clouds of the standardised difference between expected and observed numbers of headache in all 7 CCGs. Points lying below −3 or above +3 (blue lines) are considered statistically significant outliers.

## Data Availability

Due to patient data confidentiality and restrictions imposed by the HRA we are unable to directly share the data for this study. Anyone wishing to access this data must apply to do so through the HRA by completing an IRAS application.

## Acknowledgements

This research was supported by the NIHR Lancashire Clinical Research Facility. The views expressed are those of the authors and not necessarily those of the NHS, the NIHR, or the Department of Health.

## Supporting information

**S1 File. Supporting information**. Supporting information regarding variable selection and identification of potential multicolliniarity.

