## Supplementary figures and images for "Identifying variation in GP referral rates: an observational study of outpatient headache referrals"

### Supplemental Figure 1

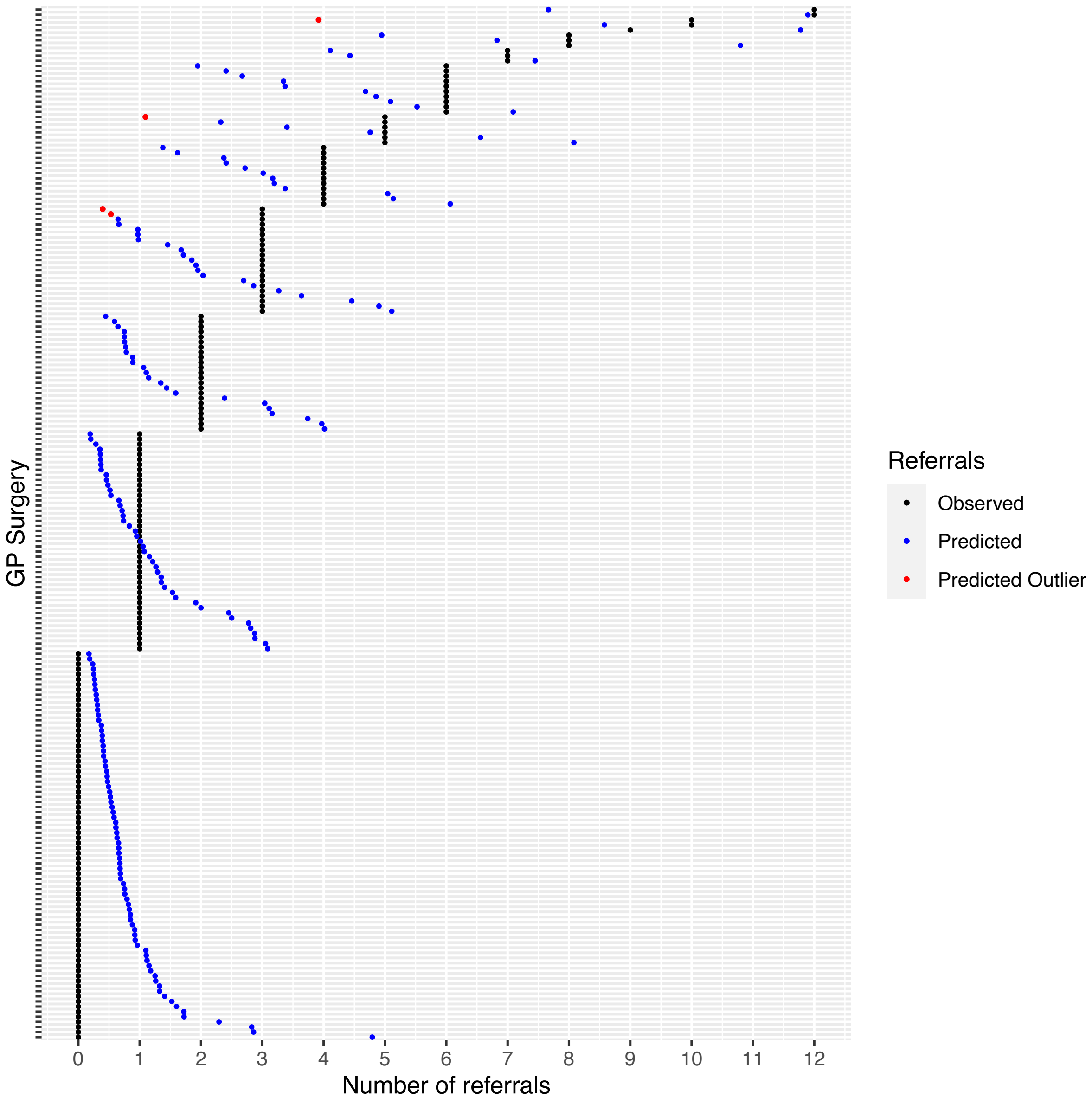
