## Supplementary text for "Identifying variation in GP referral rates: an observational study of outpatient headache referrals"

**Supporting Information**

**Identification of multicollinearity**

Data analysis was undertaken to identify potential associations and correlations between variables. Variables were tested for correlation using Pearson’s correlation coefficient. Any pair of variables found to have a Pearson’s coefficient of over 0.7 would be further tested by comparing two simple models of each variable with the outcome of interest (number of referrals for headache) and compared using the Akaike Information Criterion (AIC) to determine which one provided the best fit. The variable providing the best fit would be retained for inclusion in the main model and the less well-fitting variable discarded.

No highly correlated explanatory variables were found.

**Variable selection**

Once the best type of regression model had been selected and variables tested for multicollinearity, backwards selection was used to test models including different explanatory variables. This initially resulted in a model including only the number of other neurology referrals as an explanatory variable. However, when testing this one-variable model against the full model, the model including only other neurology referrals was found not to provide a statistically significantly better fit that the full model. Therefore, the full model was chosen in order to retain as much information about the variables as possible.

**Observed vs predicted referrals**

**Fig S1. Observed and predicted numbers of headache referrals.** Each black dot represents the observed number of headache referrals over the study period. Each blue dot represents the raw number of referrals as predicted by the poisson model. The red dots show which surgeries were determined to have referred significantly unexpected numbers of referrals.

We can clearly see the pattern of the number of observed referrals during the study period, with the largest group of surgeries referring zero patients during the study and a general trend of decreasing numbers referring larger numbers of patients. We can also see that as the groups get smaller the predictions become more difficult to make as the model has less information to work from.
